# Racial disparities in the prevalence of vaccine and non-vaccine HPV types and multiple HPV infection between Asia and Africa: A systematic review

**DOI:** 10.1101/2020.11.02.20224857

**Authors:** Jude Ogechukwu Okoye, Simon Imakwu Okekpa, Chiemeka Franklin Chukwukelu, Ifeoma Nora Onyekachi-Umeh, Anthony Ajuluchukwu Ngokere

## Abstract

**Background:** Cervical Cancer is the 6^th^ most common and 3^rd^ most deadly cancer among women. Despite the fact that majority of the countries in Asia and Africa have similar economy and low life expectancy, the mean age standardized incidence rate (ASIR) of cervical cancer is substantially higher in Africa than Asia. Thus, this study aimed to identify the correlates of the higher ASIR rates in Africa relative to Asia.

**Methods:** Peer-reviewed articles published between 2004 and 2017 were selected using the PRISMA standard. Sources of articles include Google Scholar, Scopus, PubMed Central, and EMBASE. Search keywords included: HPV genotypes, cervical cancer, HPV vaccine, and multiple infection in Africa and Asia.

**Result:** A total of 29 and 17 full-length articles were selected from Africa and Asia respectively. Based on estimates in the general population, the incidence of high-risk HPV (hrHPV) types in Africa and Asia was 3.5 and 1.0 respectively. The prevalence of HPV infection was higher in Africa than in Asia (p< 0.001). The prevalence of HPV infection between 2004-2009 and 2010-2017 decreased in Africa but increased in Asia. More so, the prevalence of multiple HPV and non-vaccine HPV infection were higher in Africa than Asia (p< 0.001). The prevalent HPV types in Africa were HPV16, HPV18, and HPV52, while that of Asia were HPV16, HPV52, and HPV58, in descending order of prevalence. This study revealed that nonavalent HPV vaccine could prevent the development of 69.3% and 83.2% of HPV associated cervical abnormalities in Africa and Asia, respectively.

**Conclusion:** This study revealed higher prevalence of HPV infection and multiple HPV infection in Africa compared with Asia, which could be responsible for the higher ASIR in Africa. It suggests that nonavalent vaccination including cervical screening using Pap smear could prevent over 90% of the cervical abnormalities in Africa.

## Introduction

Cervical cancer is the 9^th^ most common cancer in world, and the 6^th^ most common and 3^rd^ most deadly cancer among women [1]. Studies show a minor reduction in age standardized incidence rate (ASIR per 100,000) from 14.5 to 13.1 between 2017 and 2018, and a minor increase in the age standardized mortality rate (ASMR per 100,000) from 6.1 to 6.9 within the same period [1,2]. Despite the fact that majority of the countries in Asia and Africa have similar economy (less developed), low life expectancy and high mortality-to-incidence rate [3], the mean ASIR/ASMR of cervical cancer was higher in Africa (29.4/19.8) than in Asia (11.3/6.2) as of 2018 [2]. The findings of Canfell et. al. suggest that the high mortality rate among cervical cancer patients in Africa is associated with low access rate to treatment [4]. However the reason for the high incidence rate of the disease in Africa remains unknown. As of 2018, the ASIR/ASMR varies: 43.1/20.0, 40.1/30.0, 29.6/23.0/, 26.8/21.1, and 7.2/5.1 in Southern, Eastern, Western, central, and Northern Africa, respectively [2]. This shows that Northern Africa had the least ASIR/ASMR in Africa, yet these values are higher than that of Western Asia, the least affected Asian sub-region (4.1/2.5) [2]. Of note also, the ratio of cervical cancer attributable to HPV in Sub-Saharan Africa and Northern Africa/Western Asia is 9.3:1 [5]. Abryn et. al. and Martel et. al. attributed the low ASIR in Northern Africa and Western Asia to low prevalence of HPV [2,5]. They did not offer any reasons for the variation in ASIR of cervical cancer and prevalence. We however hypothesize that this could be related to the peculiar HPV subtypes found in Africa and the type of vaccines adopted in the African sub-regions.

Commonly used HPV vaccines include bivalent (HPV16/18), quadrivalent (HPV6/11/16/18) and nonavalent vaccines (Gardasil 9; 6/11/16/18/31/33/45/52/58) [6]. The first two vaccines are widely used in Africa but the nonavalent vaccine which has a global estimated efficacy of 87-89.5% in prevention of cervical cancer worldwide, is yet to be widely adopted in Africa [5,7]. Barriers to implementation of national vaccination programs in African countries include inadequate infrastructure and finances, limited health worker training, vaccine cost, and cold chain capacity constraints [8]. Even if nonavalent vaccine is widely distributed in Africa, majority of the countries in sub-Saharan Africa would not reach HPV elimination by vaccination alone. This is because, if HPV vaccination were to eradicate HPV types 6, 11, 16, 18, 31, 33, 45, 52, and 58, the incidence of cervical cancer will still be > 4/100,000 due to other non-vaccine high risk HPV (hrHPV) types that are not currently covered [9]. Furthermore, the effectiveness of this vaccine will depend on the rate of HIV infection in the target population which in turn drives multiple HPV infection. In Asia, the prevalence of multiple HPV infection in normal and abnormal cervix is 5.1-25.2% and 10.6-33.3%, respectively [10–12] whereas in Africa, this is 3.9%-15.8% and 22.9-35.7%, respectively [13–15]. This underscores the fact that multiple HPV infection limits the efficacy of available vaccine to prevent the development of cervical cancer. This review aimed at identifying the prevalent HPV types in Asia and Africa both in the general population and among women with cervical abnormalities. It also assesses the paradigm shift in the prevalence of HPV infection in Africa and Asia between two timelines; 2004-2009 and 2010-2017. It determined the prevalence of HPV infection preventable by available vaccines.

## Materials and Methods

Peer-reviewed articles published in Africa (n= 29) and Asia (n= 17) between 2004 and 2017 were selected and screened using the PRISMA standard (figure1) [16,17]. Sources of articles include Google Scholar, Scopus, PubMed Central, and EMBASE. Search keywords included “prevalence or frequency of HPV types among women Asia and Africa”, “distribution of HPV types among women with and without cervical cancer in Africa and Asia”, “vaccine and non-vaccine HPV types in Africa and Asia”, cervical cancer attributed to HPV infection in Africa and Asia”, and cervical cancer related mortality in Africa and Asia. Inclusion criteria included: Studies carried out between 2004 and 2017, studies with frequency of HPV infection, must be full-length articles and involve cervical cancer, and the articles must involve Africa and Asia. Exclusion criteria: Articles not written in English, abstracts, non-full-length article, and articles without specific frequency of HPV types, articles not involving Africa and Asia as well as articles not involving cervical cancer. Chi-square (X^2^) analysis was used to calculate the difference in HPV types between Africa and Asia (in GraphPad Prism, version 6.0). Significance was set at p< 0.05)

**Figure 1:**
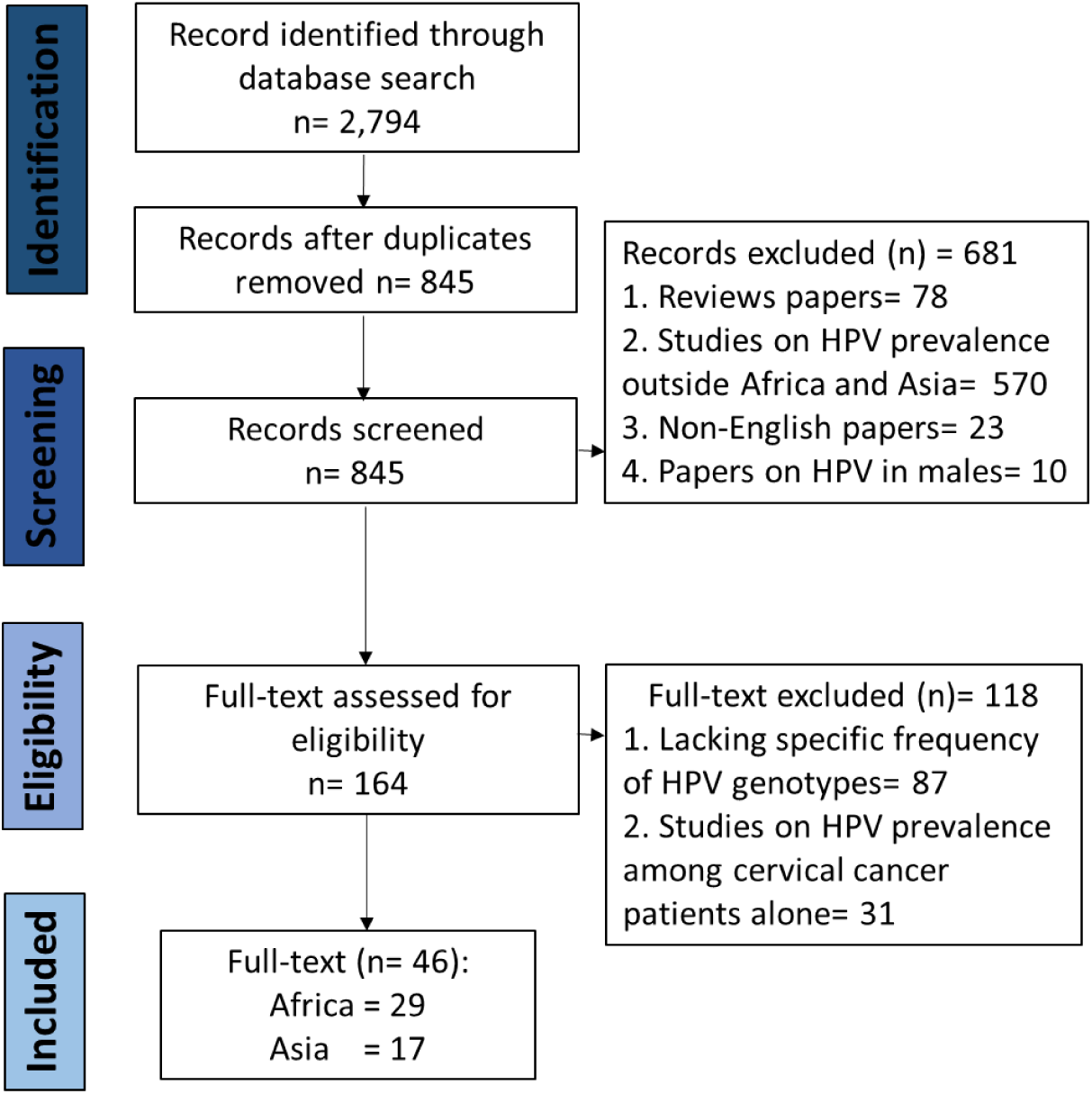
PRISMA flow Diagram on prevalence of HPV genotypes in Africa and Asia. Figure 1: Literature search and full text selection based on the PRISMA standard

## Result

Based on the inclusion criteria and according to World Health Organization classification of geographic regions, West Africa had the highest number of participants (7,313; n= 14 studies), followed by East Africa (7,240; n= 8 studies), Central Africa (2,167; n= 2 studies), North Africa (855, n= 3 studies), South Africa (153; n= 1 study). One mixed study involving Tanzania and South Africa (with 194 participants) was also included. Based on estimates in the general population, the incidence of nonavalent HPV types in Africa and Asia was 3.5 and 1.0 per 100,000, respectively. Studies in Africa revealed that HPV16, HPV18 and HPV52 ranked first or second in 62.1% (18/29), 20.7% (6/29) and 24.1% (7/29), respectively whereas studies in Asia revealed that they ranked first or second in 94.1% (16/17), 11.8% (2/17), and 41.2% (7/17), respectively. Interestingly, HPV35 ranked first and second in 17% (5/29) and 20.7% (6/29) in studies in Africa but it only ranked fifth in one study from Asia (5.9%; 1/17, table 1a and 1b). Higher prevalence of HPV and multiple infections were observed in Africa than in Asia, both in the general population and among women with cervical abnormalities (p< 0.001; tables 2–4).

**Table 1a:** Specific distribution of studies, timeline, sample size and prevalence of HPV DNA and multiple infections in Africa

| Author(s) | Location | Duration<br>of Study | AR/MA<br>years | Cases<br>N | Any HPV+<br>n (%) | Multiple<br>HPV | Order of Prev.<br>HR HPV |
| --- | --- | --- | --- | --- | --- | --- | --- |
| Obiri-Yeboah et al. [18] | Ghana | 2017 | ≤29-60<sup>+</sup>/44 | 329 | 192 (58.4) | 113 (34.3) | 35,58,52,18,56 |
| Yakub et al. [29]a | Northern Nigeria | 2016-2017 | 20-50/NA | 220 | 119 (54.1) | 25 (11.4) | 35,16,45,33,18 |
| Mutombo et al. [30] | D.R. Congo | 2015-2017 | 25-60<sup>+</sup>/46 | 1846 | 521 (28.2) | 54 (2.9) | 56,52,53,35,45 |
| Ghedira et al. [31] | Tunisia | 2016 | 17-73/NA | 471 | 160 (34.0) | 5 (1.1) | 16,31,58,66,56 |
| Ndizeye et al. [15] | Burundi | 2013/2016 | 17-65/38 | 600 | 181 (30.2) | 64 (10.7) | 16,18,33,58,31 |
| Nchome et al. [32] | Tanzania | 2015-2016 | 25-60/NA | 3416 | 644 (18.9) | NA | 52,16,58,18,35 |
| Vassilakos et al. [33] | Madagascar | 2015 | 30-65/NA | 403 | 209 (51.9) | 143 (35.4) | 53,68,52,35,16/33 |
| Marengo et al. [34] | Zimbabwe | 2015 | 18-83/39 | 136 | 70 (51.5) | 26 (19.1) | 18,52,16,58,51 |
| Belglaiaa et al. [35] | Morocco | 2014-2015 | 18-76/NA | 232 | 52 (22.4) | 34 (14.7) | 16,53,18,52,31/33/56 |
| Nyasenu et al. [36] | Togo | 2014-2015 | 20-50<sup>+</sup>/NA | 324 | 30 (9.3) | 7 (2.2) | 18,45,16,68,35/52/53 |
| Menon et al. [37] | Kenya | 2009-2015 | 18-61/28 | 616 | 355 (57.6) | 202 (32.8) | 16,53,52,56,58 |
| Krings et al. [38] | Ghana | 2014 | 18-65/31 | 1943 | 903 (46.5) | 188 (9.7) | 16,52,35,59,66 |
| Ezechi et al. [39] | Southern Nigeria | 2014 | 18-81/NA | 515 | 101 (19.6) | 28 (5.4) | 16,35,58,31,18/52 |
| Youssef et al. [40] | Egypt | 2013-2014 | 20-40/29 | 152 | 62 (40.8) | 39 (25.7) | 16,18,31,58/59,45 |
| Awua et al. [41] | Ghana | 2012-2013 | 15-65/NA | 226 | 105 (46.5) | 151 (66.8) | 16,35,58,45,18 |
| Boumba et al. [42] | Congo | 2012 | 16-72/43 | 321 | 132 (41.1) | 21 (6.5) | 16,33,18,31,35 |
| Akarolo-Anthony et al. [43] | Northern Nigeria | 2012 | 18-45<sup>+</sup>/37 | 278 | 64 (23.0) | 23 (8.3) | 35,56,58,45/59,68 |
| Reddy et al. [44]a | Malawi | 2011-2012 | 25-59/NA | 294 | 170 (57.8) | NA | 58,35,16,33/52,18 |
| Namujju et al. [22] | Uganda | 2011 | NA/23 | 1943 | 702 (36.1) | 366 (18.8) | 18,16,45,33,52 |
| Sweet et al. [45]b | Kenya | 2009-2011 | 18-50/NA | 348 | 82 (23.6) | 14 (4.0) | 52,31,16/51,35/45 |
| Dols et al. [46] | Tanzania/S. Africa | 2008-2010 | 20-73/NA | 194 | 109 (56.2) | NA | 52,16,66,33/51,35 |
| Piras et al. [14] | Benin | 2009 | 15-70/NA | 427 | 142 (33.3) | 57 (13.3) | 59,35,16,18,58 |
| Said et al. [47] | S. Africa | 2009 | NA/NA | 153 | 89 (58.2) | 40 (26.1) | 35,18,58,16,66 |
| Wolday et al. [48] | Ethiopia | 2008-2009 | 18-55+/41 | 233 | 161 (69.9) | 3 (1.3) | 16,35/45,31,56 |
| Maranga et al. [49] | Kenya | 2008-2009 | 27-68/35 | 224 | 121 (54.0) | 37 (16.5) | 56,53,52,59,58 |
| Vidal et al. [50] | Tanzania | 2008-2009 | NA/40 | 215 | 79 (36.7) | 149 (69.3) | 16,35,58,45,51 |
| Guthrie et al. [51]a | Kenya | 2007-2009 | 18-50/NA | 283 | 176 (62.2) | 122 (43.1) | 52,16/18,51,35 |
| Keita et al. [13] | Guinea | 2006-2008 | 15-64/NA | 831 | 422 (50.8) | 386 (46.5) | 16,45,52,33,35/58 |
| Okolo et al. [52] | Southern Nigeria | 2004-2006 | 15-80/NA | 313 | 126 (40.3) | 73 (23.3) | 16/35,31,56,58 |

Keys: AR; Age range, MA, mean age, Prev.; Prevalent, HR; high risk, NA; Not available, D.R.;
Democratic Republic, S. Africa; South Africa, a; HIV infected, commercial Sex Workers

**Table 1b:** Specific distribution of studies, timeline, sample size and prevalence of HPV DNA and multiple infections in Asia

| Author(s) | Location | Duration of Study | AR/MA | Cases N | Any HPV+ n (%) | Multiple HPV | Order of Prev. HR HPV |
| --- | --- | --- | --- | --- | --- | --- | --- |
| Zhang et al. [53] | East China | 2016-2017 | 18-96/44 | 2612 | 1296 (49.6) | 346 (13.2) | 16,52,58,18,53 |
| Ge et al. [11] | East China | 2016-2017 | 16-85/NA | 65613 | 10185 (15.5) | 2242 (3.4) | 16,58,52,53,39 |
| Thapa et al. [54] | Nepal | 2016-2017 | 20-65/32 | 998 | 197 (19.7) | 61 (6.1) | 16,39,58,33,51 |
| Tan et al. [55] | Malaysia | 2012-2016 | 28-77/49 | 394 | 154 (39.1) | 13 (3.3) | 16,18,58,33,31 |
| Kesheh and Keyvani [56] | Iran | 2011-2016 | 13-74/NA | 8351 | 3841 (46.0) | 1620(19.4) | 16,53,52,51,66 |
| Van et al. [57] | Vietnam | 2015 | 18-49/NA | 400 | 38 (9.5) | 14 (3.5) | 16,18,58/59,35/51/52/56 |
| Al-Lawati et al. [12] | Oman | 2014-2015 | 18-68/NA | 258 | 46 (17.8) | 11 (4.3) | 82,68,18/53/56 |
| Li et al. [58] | SW China | 2011-2015 | 17-84/36 | 28375 | 3681 (13.0) | 664 (2.3) | 52,16,58,18,56 |
| Zhong et al. [59] | SE China | 2010-2015 | 16-77/36 | 71435 | 16065 (22.5) | 5354(7.5) | 16,52,58,33,18 |
| Zhang et al. [60] | NE China | 2012-2014 | 25-65/39 | 34587 | 4198 (12.1) | 676 (2.0) | 16,58,52,53,33 |
| Zeng et al. [61] | NW China | 2011-2014 | <30-50+/NA | 51345 | 13349 (26.0) | 3382(6.6) | 16,58,56,39,18 |
| Bansal et al. [62] | Qatar | 2012-2013 | 16-84/41 | 3008 | 182 (6.1) | 6 (0.2) | 16,56,59,18/31/45 |
| Wang et al. [63] | China | 2012 | NA | 9641 | 3424 (35.5) | 486 (5.0) | 16,52,58,59,56 |
| Kantathavorn et al. [64] | Thailand | 2011-2012 | 20-70/44 | 5906 | 890 (15.1) | NA | 52,16,51,53/58 |
| Park et al. [65] | Korea | 2009-2012 | 39-58/NA | 1998 | 302 (15.1) | 40 (2.0) | 16,18,51,58,52 |
| Miyashita et al. [66]b | Philippines | 2006 | 18-40/24 | 369 | 211 (57.2) | 94 (25.5) | 52,16,66,45,59 |
| Vet et al. [67] | Indonesia | 2004-2006 | 15-70/NA | 2686 | 305 (11.4) | 63 (2.3) | 52,16,18,39,51 |

Keys: AR; age range, MA; mean age, HR; high risk, NW; North-Western, SW; South Western, NE; North East, NA; Not available, b; Commercial Sex Workers

**Table 2:** Prevalence comparison of HPV genotypes between Africa and Asia population

| Variable | Africa |  | Asia |  | Africa vs Asia |  |
| --- | --- | --- | --- | --- | --- | --- |
|  | Cases<br>(N) | HPV+<br>n (%) | Cases<br>(N) | HPV+<br>n (%) | % difference<br>(Rank) | X<sup>2</sup> (p) |
| **Any HPV** | 17486 | 6541 (37.4) | 287976 | 58364 (20.3) | 17.1 | <0.001 |
| **Multiple** | 13582 | 2482 (18.3) | 282070 | 16274 (5.7) | 12.6 | <0.001 |
| **Vaccine HR** |  |  |  |  |  |  |
| HPV16 | 17486 | 1396 (8.0) | 287976 | 9413 (3.2) | 4.8 (1) | <0.001 |
| HPV18 | 17486 | 927 (5.3) | 287976 | 2765 (1.0) | 4.3 (3) | <0.001 |
| HPV31 | 17486 | 669 (3.8) | 287976 | 1294 (0.4) | 3.4 (4) | <0.001 |
| HPV33 | 17253 | 439 (2.5) | 287976 | 1952 (0.7) | 1.8 (9) | <0.001 |
| HPV45 | 17165 | 669 (3.9) | 287976 | 534 (1.9) | 2.0 (8) | <0.001 |
| HPV52 | 17165 | 857 (5.0) | 287976 | 7686 (2.7) | 2.3 (7) | <0.001 |
| HPV58 | 17486 | 641 (3.7) | 287976 | 5878 (2.0) | 1.7 (10) | <0.001 |
| **Other HR** |  |  |  |  |  |  |
| HPV35 | 17486 | 801 (4.6) | 287976 | 578 (0.2) | 4.4 (2) | <0.001 |
| HPV39 | 17165 | 247 (1.4) | 287976 | 2679 (0.9) | 0.5 (16) | <0.001 |
| HPV51 | 16932 | 402 (2.4) | 287976 | 2293 (0.8) | 1.6 (12) | <0.001 |
| HPV53 | 11928 | 312 (2.6) | 276270 | 2098 (0.8) | 2.8 (5) | <0.001 |
| HPV56 | 17486 | 388 (2.2) | 287976 | 2391 (0.8) | 1.4 (14) | <0.001 |
| HPV59 | 16852 | 370 (2.2) | 287976 | 1537 (0.5) | 1.7 (10) | <0.001 |
| HPV66 | 12927 | 408 (3.2) | 278668 | 1243 (0.4) | 2.8 (5) | <0.001 |
| HPV68 | 16738 | 373 (2.2) | 284968 | 1608 (0.6) | 1.6 (12) | <0.001 |
| HPV82 | 9045 | 158 (1.7) | 101345 | 607 (0.6) | 1.1 (15) | <0.001 |
| **LR Type** |  |  |  |  |  |  |
| HPV6 | 9265 | 186 (2.0) | 180707 | 4188 (2.3) | 0.3 (17) | 0.057 |
| HPV11 | 8241 | 132 (1.6) | 180707 | 2404 (1.3) | 0.3 (17) | 0.034 |

**Table 3:** Frequency of HPV genotype among women with abnormal cervix

| Variable | Africa |  | Asia |  | Africa vs Asia | Africa vs Asia |
| --- | --- | --- | --- | --- | --- | --- |
| Abnormal | Cases | HPV+ | Cases | HPV+ | Diff. in | X<sup>2</sup> (p) |
| Cervix | (N) | n (%) | (N) | n (%) | % (Rank) |  |
| **Any HPV** | 1035 | 815 (78.7) | 961 | 820 (85.3) | 6.6 | <0.001 |
| **Multiple** | 544 | 168 (30.9) | 757 | 159 (21.0) | 9.9 | <0.001 |
| **Vaccine** |  |  |  |  |  |  |
| **HR** |  |  |  |  |  |  |
| HPV16 | 1035 | 366 (35.3) | 961 | 358 (37.3) | 2.0 (15) | 0.402 |
| HPV18 | 1035 | 108 (10.4) | 961 | 69 (7.2) | 3.2 (9) | 0.012 |
| HPV31 | 1035 | 78 (7.5) | 961 | 25 (2.6) | 4.9 (4) | <0.001 |
| HPV33 | 972 | 52 (5.3) | 961 | 71 (7.4) | 2.1 (14) | 0.076 |
| HPV45 | 1035 | 84 (8.1) | 961 | 12 (1.2) | 6.9 (3) | <0.001 |
| HPV52 | 960 | 136 (14.2) | 961 | 156 (16.2) | 2.0 (15) | 0.227 |
| HPV58 | 1035 | 104 (10.0) | 961 | 141 (14.7) | 4.7 (5) | 0.002 |
| **Other HR** |  |  |  |  |  |  |
| HPV35 | 960 | 119 (12.4) | 961 | 15 (1.6) | 10.8 (1) | <0.001 |
| HPV39 | 960 | 25 (2.6) | 961 | 22 (2.3) | 0.3 (17) | 0.661 |
| HPV51 | 960 | 62 (6.5) | 961 | 28 (2.9) | 3.6 (8) | <0.001 |
| HPV53 | 742 | 23 (3.1) | 961 | 32 (3.3) | 0.2 (18) | 0.890 |
| HPV56 | 972 | 47 (4.8) | 961 | 16 (1.7) | 3.1 (10) | <0.001 |
| HPV59 | 960 | 33 (3.4) | 961 | 20 (2.1) | 2.3 (13) | 0.072 |
| HPV66 | 867 | 58 (6.7) | 961 | 23 (2.3) | 4.4 (6) | <0.001 |
| HPV68 | 897 | 34 (3.8) | 961 | 9 (0.9) | 2.9 (11) | <0.001 |
| HPV82 | 769 | 27 (3.5) | 757 | 7 (0.9) | 2.6 (12) | 0.001 |
| **LR Type** |  |  |  |  |  |  |
| HPV6 | 245 | 20 (8.2) | 757 | 9 (1.2) | 7.0 (2) | <0.001 |
| HPV11 | 308 | 16 (5.2) | 757 | 10 (1.3) | 3.9 (7) | 0.001 |

**Table 4:**
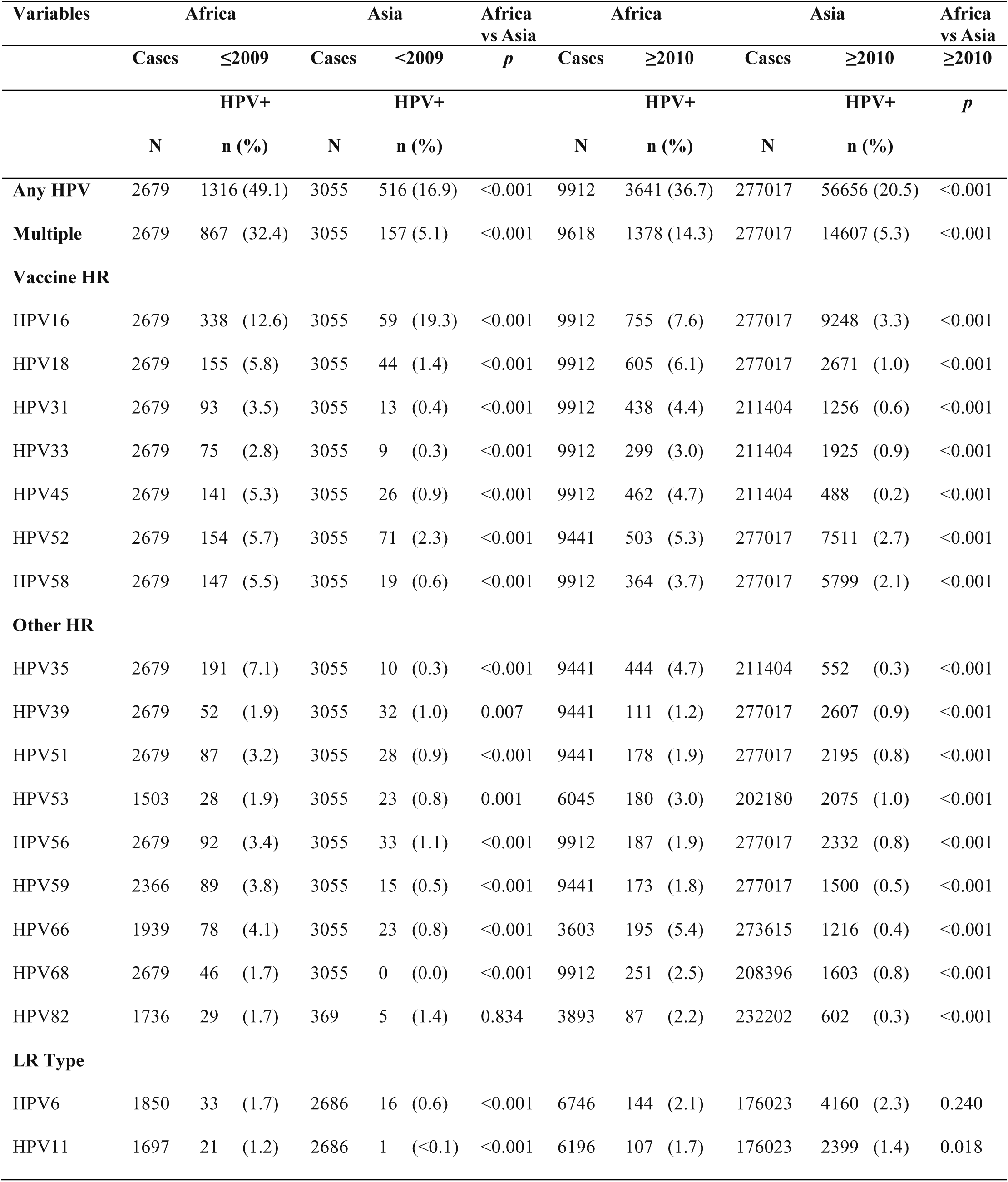
Prevalence comparison of HPV genotypes in African and Asian timeline

### Prevalence of HPV types in multiple infection

Considering the general population, significant differences in the prevalence of HPV types were observed between Africa and Asia, except for HPV6 (p< 0.001). High differences were observed between the two continents with regards to HPV16, HPV35, HPV18, HPV31 and HPV53/66 prevalence, in descending order of rank (table 2). There were no observed differences between African and Asian studies with respect to vaccine hrHPV (p> 0.05) whereas, significant differences were observed between African and Asian studies with respect to non-vaccine hrHPV (p< 0.001). lrHPV showed no differences between African and Asian studies (p> 0.05; figure 2a). The first five HPV types with the highest involvement in multiple infection in Africa were HPV6, HPV16, HPV11, HPV35 and HPV45 while that of Asia were HPV16, HPV58, HPV33, HPV52 and HPV53, in order of descending rank (figure 2b).

**Figure 2:**
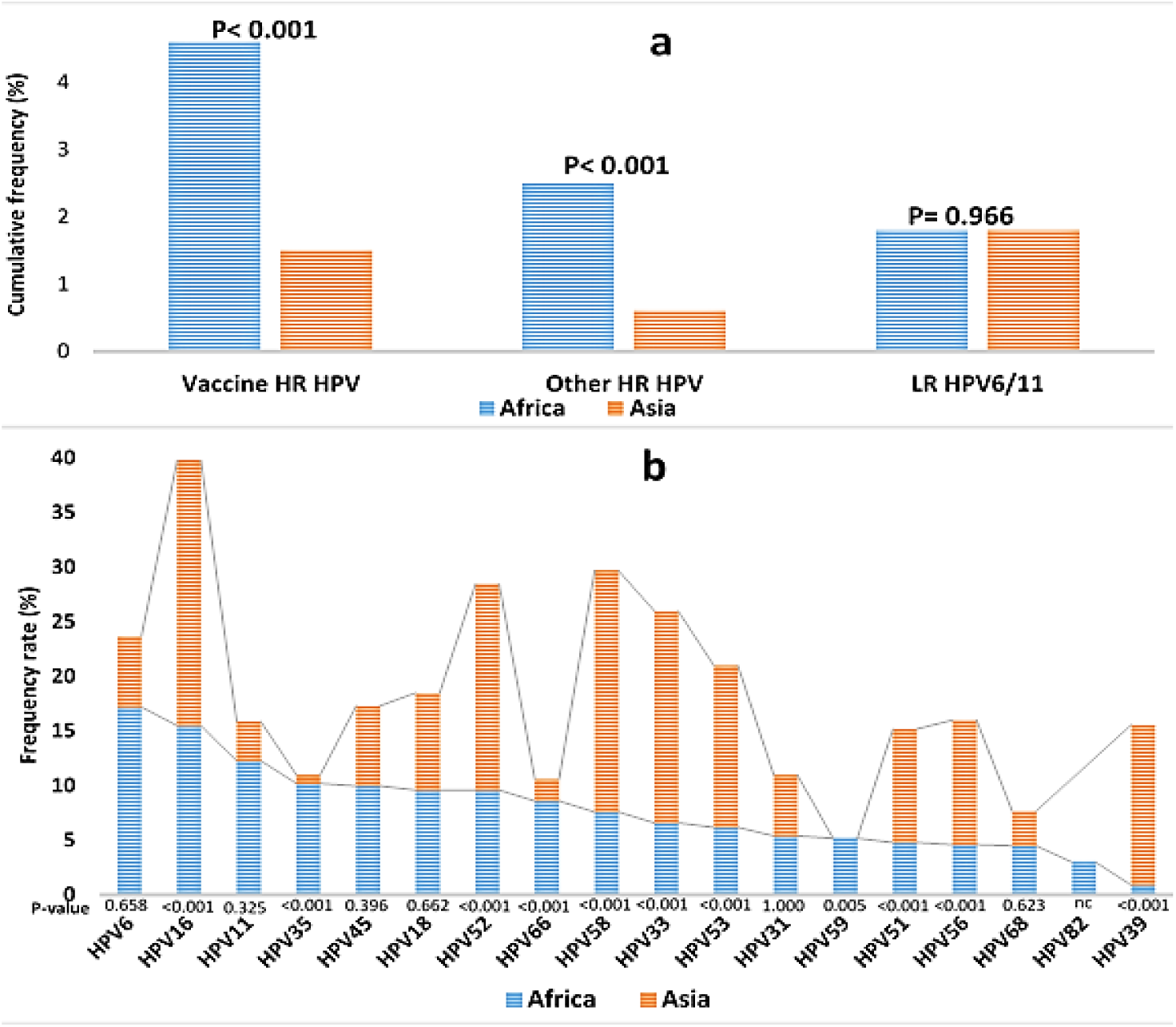
Frequency of HPV genotypes in the general population of Africa and Asia. Figure 2a: Higher prevalence of vaccine and non-vaccine HPV types were found in African than in Asia (p< 0.001) while similar prevalence of lrHPV types was found in Africa and Asia (p> 0.05). Figure 2b: In terms of multiple infection, significant differences were observed between Africa and Asia with regards to HPV58, HPV39, HPV33, HPV35, HPV52, HPV16, HPV53, HPV56, HPV66, HPV51, HPV59, HPV45, HPV68, HPV18 and HPV31, in descending order of rank. No comparison (nc) was carried out in respect to HPV82 due to the fact it was not investigated in the selected Asian studies used for plotting this graph.

### Prevalence of HPV types among women with cervical abnormalities

The prevalent HPV types in Africa were HPV16, HPV52, HPV35, HPV18 and HPV58 while the prevalent HPV types in Asia were HPV16, HPV52, HPV58, HPV33, and HPV53, in descending order of rank. High significant differences were observed between Africa and Asia with regards to HPV35, HPV6, HPV45, HPV31, HPV58, HPV66 and HPV68, in descending order of rank (table 3). Result showed a higher prevalence of hrHPV and lrHPV in Africa than in Asia (p< 0.001; figure 3a). It also shows that nonavalent HPV vaccine could prevent the development of 69.3% and 83.2% of HPV attributable cervical abnormalities in Africa and Asia, respectively whereas bivalent and quadrivalent vaccines could prevent the development of about 30% and 40% of HPV associated cervical abnormalities in Africa and Asia, respectively (figure 3b).

**Figure 3:**
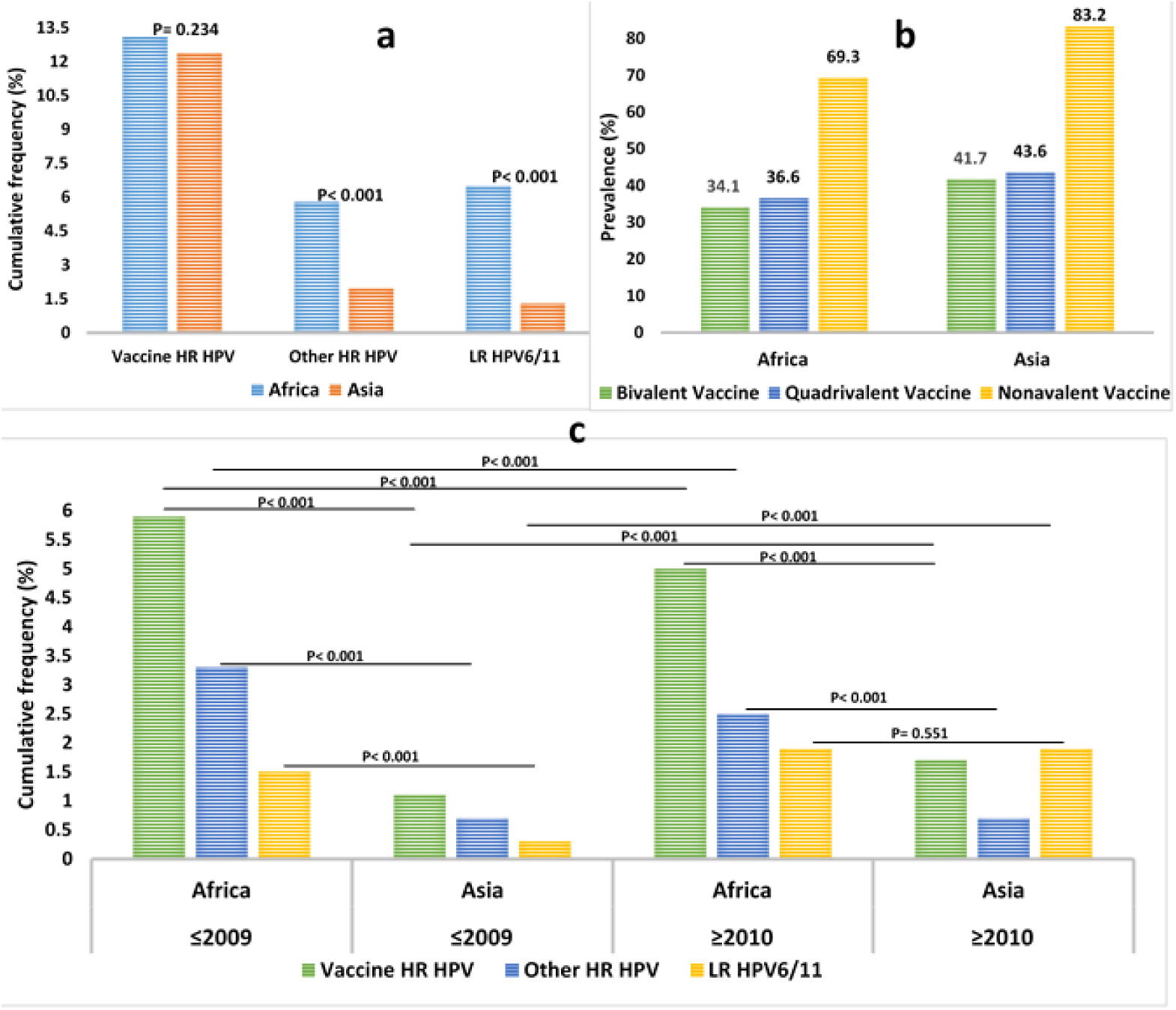
a) Frequency comparison of HPV genotypes in African and Asian women irrespective of cervical status, b) Prevalence of HPV-associated cervical abnormalities preventable by available vaccines and c) Cumulative frequency comparison of vaccine and Non-vaccine HPV between Africa and Asia with regards to year. Figure 3: No significant difference was observed between Africa and Asia studies in terms of vaccine hrHPV genotypes. However, significance differences were observed between Africa and Asian studies in terms of non-vaccine other hrHPV types and lrHPV16/11 (p< 0.001; figure 3a). Figure 3b shows that quadrivalent vaccine may have mild edge over bivalent vaccine in preventing the development of cervical abnormalities both in Africa and Asia while nonavalent vaccine could exert almost twice the effect of bivalent and quadrivalent vaccines in forestalling the development of cervical abnormalities. Figure 3c: Cumulatively, as of 2009 and 2017, the prevalence of vaccine and non-vaccine hrHPV were significantly higher in Africa than in Asia (p< 0.001). As of 2009, the prevalence of lrHPV was significantly higher in Africa than in Asia (p< 0.001) while the prevalence of the viruses were similar between both Continents in 2017 at p> 0.05.

### Difference in HPV prevalence between 2004-2009 and 2010-2017

There was decreased HPV infection from 2004-2009 in Africa whereas Asia witnessed increase in HPV infection from 2010-2017 (table4). In 2009, there was a significant difference in the prevalence of HPV types in Africa and Asia (p≤ 0.01), excluding HPV82 (p> 0.05). More so, the difference in the prevalence of hrHPV types between Africa and Asia in 2017 was significant (p< 0.001). No significant difference was observed between the two continents in terms of the prevalence of HPV6 and HPV11 (p> 0.05 and p< 0.05), respectively (table 4). In 2004-2009, the prevalence of vaccine (5.9% vs 1.1%) and non-vaccine HPV (3.3% vs 0.7%) were higher in Africa than in Asia. Similarly, in 2010-2017, the prevalence of vaccine (5% vs 1.7%) and non-vaccine HPV (2.5% vs 0.7%) were higher in Africa than in Asia (p< 0.001; figure 3c). Furthermore, in Africa and Asia, the prevalence of multiple HPV infection decreased and increased by 18.1% and 0.3% from 2004-2009 to 2010-2017, respectively. Both in Africa and Asia, the prevalence of HPV16, HPV56, HPV51, HPV39 and HPV45 decreased from 2004 to 2017 (5% vs 16.0%, 1.5% vs 0.3%, 1.3% vs 0.1%, 0.7% vs 0.1%, and 0.6% vs 0.7%, respectively) while the prevalence of HPV53, HPV31, HPV68 and HPV33 (1.1% vs 0.2%, 0.9% vs 0.2%, and 0.3% vs 0.8%, and 0.2% vs 0.6%, respectively) increased from 2004 to 2017. This suggests that among hrHPVs the prevalence of HPV16 decreased the most whereas HPV53 increased the most. In Africa, the prevalence of HPV35, HPV59, HPV58, and HPV52 decreased 2.4%, 2.0%, 1.8%, and 0.4% from 2004-2009 to 2010-2017 while the prevalence of HPV66 and HPV18 increased by 1.3%, and 0.3%, respectively. In Asia, the prevalence of HPV82 and HPV18/66 decreased from 2004-2009 to 2010-2017 by 1.1% and 0.4%, respectively. Conversely, the prevalence of HPV58 and HPV52 increased by 1.5%, and 0.4%, respectively while the prevalence of HPV35 and HPV59 remained unchanged (table 4).

### Prevalence of HPV types in the African sub-regions

The highest and lowest prevalence of HPV infection as well as highest and lowest multiple infections were found in South Africa and Central Africa respectively. South Africa had the highest prevalence of HPV18, HPV45, HPV58, HPV35, HPV39, HPV51, and HPV59 while West Africa had the highest prevalence of HPV52, HPV53, HPV56, HPV66, and HPV82. East Africa had the highest prevalence of HPV31, HPV33, and HPV11 while North Africa had the highest prevalence of HPV16 and HPV6. Central Africa had the highest prevalence of HPV68. North Africa had the lowest prevalence of HPV45, HPV52, HPV58, HPV35, HPV66, HPV68, and HPV82 (table 5). Result showed that the prevalence of non-vaccine hrHPV types were lower in the North and East Africa (figure 4a) while the prevalence of HPV preventable by nonavalent vaccine was higher in both regions than other African sub-regions (figure 4b).

**Table 5:** Prevalence of some high- and low-risk HPV genotypes in sub-regions of Africa

**Tables 5: Prevalence of some high- and low-risk HPV genotypes in sub-regions of Africa**
| Variables | North Africa |  | East Africa |  | Central Africa |  | West Africa |  | South Africa |  |
| --- | --- | --- | --- | --- | --- | --- | --- | --- | --- | --- |
|  | Cases | HPV+ | Cases | HPV+ | Cases | HPV+ | Cases | HPV+ | Cases | HPV+ |
|  | N | n (%) | N | n (%) | N | n (%) | N | n (%) | N | n= (%) |
| **Any HPV** | 855 | 274 (32.0) | 7240 | 2216 (30.6) | 2167 | 653 (30.1) | 7313 | 3286 (44.9) | 153 | 89 (58.2) |
| **Multiple** | 855 | 78 (9.1) | 3530 | 863 (24.4) | 2167 | 75 (3.5) | 7313 | 1464 (20.0) | 153 | 40 (26.1) |
| **Vaccine HR** |  |  |  |  |  |  |  |  |  |  |
| HPV16 | 855 | 97 (11.3) | 7240 | 645 (8.9) | 2167 | 93 (4.3) | 7313 | 602 (8.2) | 153 | 17 (11.1) |
| HPV18 | 855 | 27 (3.2) | 7240 | 485 (6.7) | 2167 | 53 (2.4) | 7313 | 390 (5.3) | 153 | 15 (9.8) |
| HPV31 | 855 | 21 (2.5) | 7240 | 371 (5.1) | 2167 | 50 (2.3) | 7313 | 268 (3.7) | 153 | 6 (3.9) |
| HPV33 | 855 | 13 (1.5) | 7007 | 270 (3.9) | 2167 | 29 (1.3) | 7313 | 177 (2.4) | 153 | 1 (0.7) |
| HPV45 | 855 | 9 (1.1) | 7240 | 374 (5.2) | 1846 | 47 (2.5) | 7313 | 279 (3.8) | 153 | 9 (5.9) |
| HPV52 | 384 | 10 (2.6) | 7240 | 382 (5.3) | 1846 | 66 (3.6) | 7313 | 502 (6.9) | 153 | 6 (3.9) |
| HPV58 | 855 | 19 (2.2) | 7240 | 276 (3.8) | 2167 | 73 (3.4) | 7313 | 275 (3.8) | 153 | 14 (9.1) |
| **Other HR** |  |  |  |  |  |  |  |  |  |  |
| HPV35 | 384 | 5 (1.3) | 7240 | 313 (4.3) | 2167 | 56 (2.6) | 7313 | 470 (6.4) | 153 | 14 (9.1) |
| HPV39 | 384 | 4 (1.0) | 7240 | 46 (0.6) | 1846 | 32 (1.7) | 7313 | 174 (2.4) | 153 | 13 (8.5) |
| HPV51 | 384 | 9 (2.3) | 7007 | 120 (1.7) | 1846 | 42 (2.3) | 7313 | 276 (3.8) | 153 | 8 (5.2) |
| HPV53 | 703 | 11 (1.6) | 4867 | 74 (1.5) | 1846 | 60 (3.3) | 5560 | 240 (4.3) | 153 | -- |
| HPV56 | 855 | 10 (1.2) | 7240 | 87 (1.2) | 2167 | 23 (1.1) | 7313 | 301 (4.1) | 153 | 3 (2.0) |
| HPV59 | 384 | 7 (1.8) | 7240 | 60 (0.8) | 1846 | 26 (1.4) | 7000 | 286 (4.1) | 153 | 8 (5.2) |
| HPV66 | 855 | 15 (1.8) | 4867 | 216 (4.4) | 2167 | 47 (2.2) | 5456 | 298 (5.5) | 153 | 8 (5.2) |
| HPV68 | 855 | 7 (0.8) | 7240 | 99 (1.4) | 1846 | 102 (5.5) | 6886 | 232 (3.4) | 153 | 7 (4.6) |
| HPV82 | 705 | 5 (0.7) | 4328 | 66 (1.5) | -- | -- | 4626 | 88 (1.9) | 153 | 2 (1.3) |
| **LR Type** |  |  |  |  |  |  |  |  |  |  |
| HPV6 | 855 | 26 (3.0) | 1218 | 28 (2.3) | 2167 | 19 (0.9) | 4872 | 110 (2.3) | 153 | 3 (2.0) |
| HPV11 | 855 | 13 (1.5) | 1218 | 59 (4.8) | 1846 | 8 (0.4) | 4548 | 52 (1.1) | -- | -- |

Keys: S; South, N; North, E; East, W; West, C; Central

**Figure 4:**
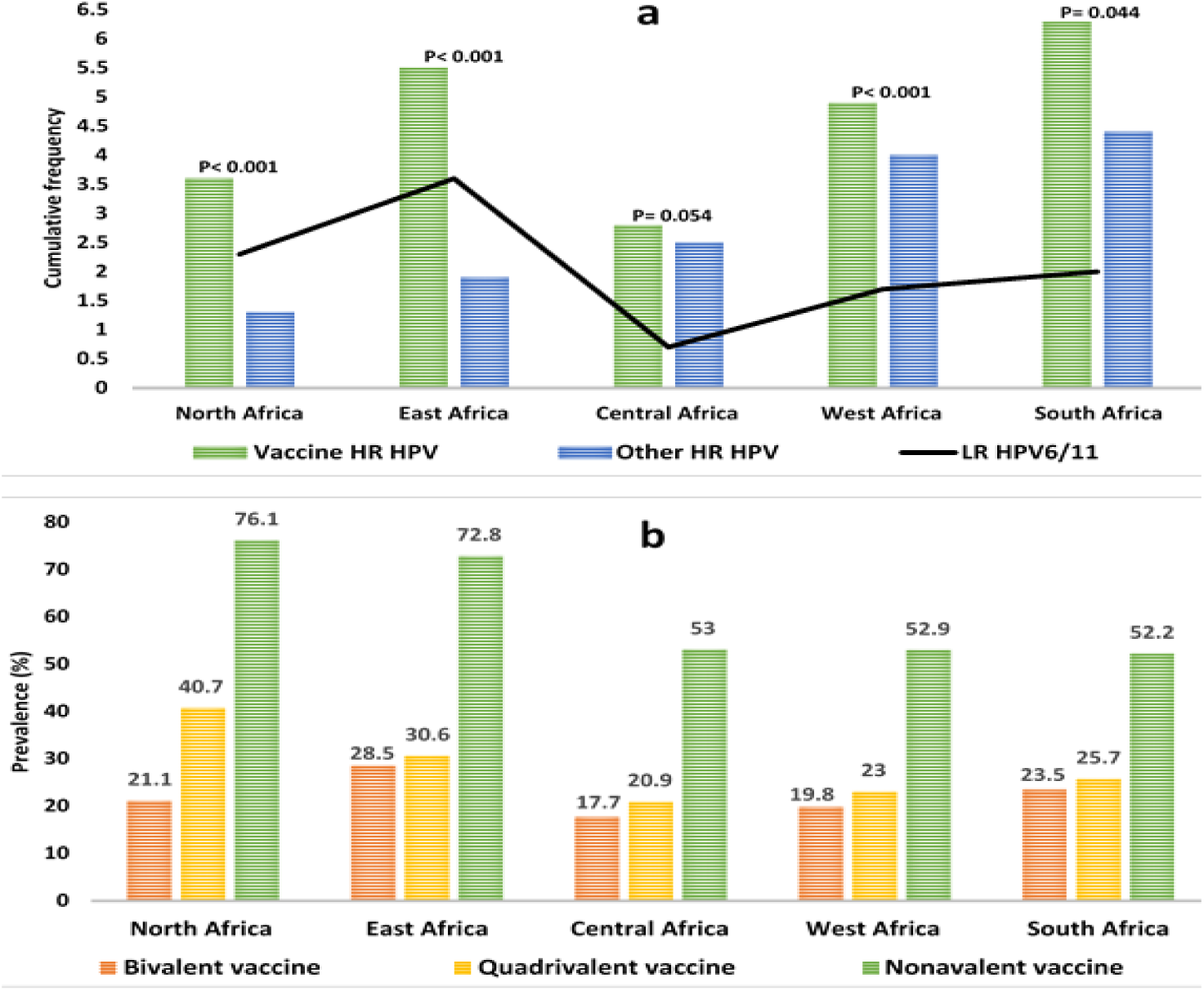
HPV genotypes and HPV preventable by HPV vaccines based on African sub-regions. Figure 4a shows that the prevalence of non-vaccine (other) hrHPV infection is highest in South Africa but lowest In North Africa. It also shows that the prevalence of vaccine hrHPV infection and lrHPV infection was higher in South Africa and East Africa, respectively while the prevalence of vaccine hrHPV and lrHPV infections were lower in central Africa. Higher differences in prevalence of vaccine and non-vaccine hrHPV types were observed in North and East African sub-regions (p< 0.001). Figure 4b shows that the prevalence of HPV infection preventable by quadrivalent and nonavalent vaccines were higher in North and East Africa than in other African sub-regions.

## Discussion

The incidence of cervical cancer in a population majorly depends on HPV and cervical cancer awareness, extent of high-risk sexual behaviour, prevalence of HPV infection and rate of HPV vaccination (2,5,18]. Due to low HPV vaccine coverage in Sub-Sahara Africa (1.2%), and lack of implementation of World Health Organization (WHO) and Center for Disease control and prevention (CDCP)’s recommendations [6,19], a high ASIR of cervical cancer is expected in Africa [2,18]. However, there is a need to indirectly assess the past and ongoing cervical cancer related interventions. There is also the need to identify major contributors to the high ASIR and ASMR in Africa for appropriate channeling of future interventions following recommendations. In the general population, this study revealed that HPV16, HPV52 and HPV58 were the most prevalent hrHPV types in Asia whereas HPV16, HPV18 and HPV52 were the most prevalent hrHPV infections in Africa. Considering the prevalence of hrHPV in abnormal cervix, this study revealed striking differences between Africa and Asia with regards to the prevalence of HPV35, HPV45, HPV31, HPV58, and HPV66. According to Hashim et al., the risk of developing CIN2+/CIN3+ among HPV16 and HPV18 positive women following a regular follow-up of 21 months are 24.4/19.9 and 16.2/10.8, respectively while the risk for women who are positive for other hrHPVs is 9.6/5.5 [20]. Based on the latter, the substantial higher prevalence of HPV16 and HPV18 in Africa relative to Asia might also be an explanation for the difference in cervical cancer ASIR between the two continents as revealed by Fitzmaurice et al. and Abryn et al. [1,2]. Considering the fact that HPV16 infection is associated with high mortality among HPV positive women [21], it could be argued that the high prevalence of HPV is linked to the high ASMR or poor prognosis among African women [1–3].

Namujju et al. stated that HPV16- and HPV18- positive women have 11-22-fold and 45-58 increased risk of acquiring other hrHPVs, respectively [22]. Thus, the higher prevalence of multiple HPV infection observed in Africa relative to Asia could be due to the higher prevalence of HPV16 and HPV18 in African women. The multiple HPV infection could be responsible for the higher ASIR in Africa than in Asia since higher prevalence of multiple infection has been reported in abnormal cervix than in normal cervix [13–15]. In addition, multiple HPV infection has been linked to increasing prevalence of human immunodeficiency virus (HIV) which in turn contributes significantly to cervical carcinogenesis [18]. Thus, it could be suggested that the high ASIR of cervical cancer in Africa when compared with Asia stems from a cascade of event beginning from non-vaccine HPV infection and high multiple infection. In this study, South Africa had the highest prevalence of any HPV and multiple HPV infection while Central Africa had the least of both parameters. The difference between the African sub-regions in terms of ASIR of cervical cancer as revealed by Martel et al. [5] could be due to high multiple HPV infections and HIV infection rate [23,24].

According to Huh et al. [25], nonavalent vaccine prevents 90% of cervical cancer worldwide. Their estimate is substantially higher than the estimate of this study (69%). This could be attributed to their study not including any participants from African countries. Cervical cancers can be attributed to a single hrHPV types or multiple types such as HPV16, HPV18, HPV31, HPV33, HPV45, HPV52, and HPV58 which are preventable by available vaccines [5]. However, the vaccine hrHPV and non-vaccine hrHPV may not be effective in preventing multiple infection mediated cervical cancer involving HPV16/35, HPV16/53, HPV16/66, except in the event of cross-protection. Pirek et al. stated that there is an increased rate of malignant transformation in women with normal cervix and multiple HPV infection (< 365days) than in women with normal cervix and single HPV infection [26], hence the need for continual screening, even among vaccinated women and immunocompromised women. Given the high rate of multiple infections in Africa, especially West and South Africa, nonavalent vaccine may offer better protection against cervical cancer than other vaccines. Additionally, the hrHPV type with the highest difference between African and Asian women diagnosed with cervical abnormalities was HPV35. Consequently, the high prevalence of HPV35 and its involvement in multiple infection may account for the higher ASIR in Africa than in Asia [1,2]. Sadly, none of the available vaccines offer protection against HPV35, the second most prevalent HPV types which accounts for 12.4% of cervical abnormalities in Africa. Since HPV35 appears to be the major correlate of cervical carcinogenesis in Africa, continual Pap smear screening is recommended.

In this study, between 2004 and 2017, the prevalence of HPV infection and multiple infection decrease by 12.4% and 18.1% in Africa while they increased by 3.5% and 0.2% in Asia, respectively. The reason for the observed decrease in Africa could majorly be due to increasing awareness. On the other hand, considering the fact that the widely distributed vaccine in Africa are bivalent and quadrivalent vaccines [5,7], the decrease could be attributed to HPV vaccination since the prevalence of HPV16 substantially decreased (5%). Despite the fact that the overall prevalence of HPV infection decreased between 2004 and 2017 in Africa, the prevalence of HPV 33, 18, 6, 11, 82, 68, 31, 53, and 66 increased by 0.2%, 0.3%, 0.4%, 0.5%, 0.5%, 0.8%, 0.9%, 1.1%, and 1.3%, respectively within the same period, majority of the HPV types (56%; 5/9) are not covered by available vaccines. This may severely impact on the prevalence of cervical cancer in Africa. The reason however for the increased prevalence of HPV in Asia from 2004-2017 is still unknown. Interestingly, in Asia and Africa, the prevalence of HPV16 decreased by 16% and 5%, respectively while the following HPV types increased by certain percentage in 2017: HPV31 (0.2% vs 0.9%), HPV53 (0.2% vs 1.1%), HPV33 (0.6% vs 0.2%), HPV68 (0.8% vs 0.7%), HPV11 (1.3% vs 0.5%), and HPV6 (1.7% vs 0.4%, respectively). Differentially, the prevalence of HPV 52 and 58 increased in Asia by 0.5%, and 1.5%, respectively while in Africa the prevalence of HPV 18, 82 and 66 increased by 0.3%, 0.5% and 1.3%, respectively (table 4). Though the prevalence of HPV 6, 11, 31, 33, 53, and 68 increased in both continents, the findings of this study suggest that if both continents adopt only nonavalent vaccines, it would still take a longer time to eradicate or significantly reduce cervical cancer in Africa than it would in Asia. This reiterates the fact that cervical cancer screening using Pap smear should still be an integral part of preventive measure in order to eliminate the disease in Africa by 2090 [9].

The prevalence of vaccine and non-vaccine hrHPV were higher in South Africa than in other African sub-region while the prevalence of vaccine and non-vaccine hrHPV were lowest in Central Africa and North Africa. This study suggests that majority, over 72%, of cervical cancer attributable to HPV in North and East Africa could be prevented by vaccine, especially by using quadrivalent and nonavalent vaccines. Since 7 out of 12 countries are currently providing HPV vaccination at no cost for girls in East Africa, there is a higher chance of eliminating cervical cancer in the sub-continent by 2090 [9,27]. The findings of this study show that though nonavalent vaccine could prevent approximately 52% of cervical cancer attributable to HPV in Central, Western and Southern Africa, a tangible number of cervical cancer will still be observed in the sub-regions due to high prevalence of non-vaccine hrHPV.

## Conclusion

This study revealed that the disparity between African and Asian women with regards to ASIR could be linked to high-risk HPV infection and multiple HPV types.. It suggests that nonavalent vaccination including cervical screening using Pap smear could prevent over 90% of the cervical abnormalities in Africa.

## Data Availability

Data are available and will be provided on request

## References

1. Fitzmaurice C, Abate D, Abbasi N, et al. Global, regional, and national cancer incidence, mortality, years of life lost, years lived with disability, and disability-adjusted life-years for 29 cancer groups, 1990 to 2017: a systematic analysis for the global burden of disease study. JAMA Oncol. 2019; 5 (12), 1749–68.

2. Arbyn M, Weiderpass E, Bruni L, et al. Estimates of incidence and mortality of cervical cancer in 2018: a worldwide analysis. The Lancet Global Health. 2020; 8 (2), e191–203.

3. Chen SL, Wang SC, Ho CJ, et al. Prostate cancer mortality-to-incidence ratios are associated with cancer care disparities in 35 countries. Sci Rep. 2017; 7 (1), 1–6.

4. Canfell K, Caruana M, Gebski V, et al. Cervical screening with primary HPV testing or cytology in a population of women in which those aged 33 years or younger had previously been offered HPV vaccination: Results of the Compass pilot randomised trial. PLoS Med. 2017; 14 (9), e1002388.

5. de Martel C, Plummer M, Vignat J, Franceschi S. Worldwide burden of cancer attributable to HPV by site, country and HPV type. Int J Cancer. 2017; 141 (4), 664–70

6. Center for Disease Control and Prevention. Morbidity and Mortality Weekly Report. Human Papillomavirus Vaccination for Adults: Updated Recommendations of the Advisory Committee on immunization Practices. Weekly. 2019; 68 (32), 698–70

7. Muñoz N, Bosch FX, de Sanjosé S, et al. Epidemiologic classification of human papillomavirus types associated with cervical cancer. N Engl J Med. 2003; 348 (6) :518–27

8. Black E, Richmond R. Prevention of Cervical Cancer in Sub-Saharan Africa: The Advantages and Challenges of HPV Vaccination. Vaccines 2018; 6 (3), 61

9. Brisson M, Kim JJ, Canfell K, et al. Impact of HPV vaccination and cervical screening on cervical cancer elimination: a comparative modelling analysis in 78 low-income and lower-middle-income countries. The Lancet. 2020; 395 (10224), 575–90.

10. Muderris T, Afsar I, Yildiz A, Varer CA. HPV genotype distribution among women with normal and abnormal cervical cytology in Turkey. Rev Esp Quimioter. 2019; 32 (6), 516–24.

11. Ge Y, Zhong S, Ren M, Ge Y, Mao Y, Cao P. Prevalence of human papillomavirus infection of 65,613 women in East China. BMC Public Health 2019; 19 (1), 178

12. Al-Lawati Z, Khamis FA, Al-Hamdani A, et al. Prevalence of human papilloma virus in Oman Genotype 82 and 68 are dominating. Int J Infect Dis. 2020; 93, 22–27

13. Keita N, Clifford GM, Koulibaly M, et al. HPV infection in women with and without cervical cancer in Conakry, Guinea. Br J Cancer. 2009; 101 (1), 202–08

14. Piras F, Piga M, Montis AD, et al.: Prevalence of Human Papillomavirus infection in women in Benin, West Africa. Virol J. 2011; 8 (1), 514.

15. Ndizeye Z, Vanden Broeck D, Lebelo RL, Bogers J, Benoy I, Van Geertruyden JP. Prevalence and genotype-specific distribution of human papillomavirus in Burundi according to HIV status and urban or rural residence and its implicationsfor control. PLoS One. 2019; 14 (6), e0209303

16. Liberati A, Altman DG, Tetzlaff J, et al. (2009). The PRISMA statement for reporting systematic reviews and meta-analyses of studies that evaluate health care interventions: explanation and elaboration. PLoS Med. 2009; 62 (10), e1–34.

17. Moher D, Liberati A, Tetzlaff J, Altman DG, Prisma Group. Preferred reporting items for systematic reviews and meta-analyses: the PRISMA statement. PLoS Med. 2009; 6 (7), e1000097.

18. Klein C, Kahesa C, Mwaiselage J, West JT, Wood C, Angeletti PC. How the Cervical Microbiota Contributes to Cervical Cancer Risk in Sub-Saharan Africa. Front Cell Infect Microbiol. 2020; 10:23.

19. World Health Organisation (WHO). Comprehensive Cervical Cancer Control: A Guide to Essential Practice; World Health Organisation: Geneva, Switzerland, 2014; ISBN 978-92 4-154895-3.

20. Hashim D, Birgit Engesæter, Gry Baadstrand Skare, Philip E. Castle, Tone Bjørge, Ameli Tropé and Mari Nygård. Real-world data on cervical cancer risk stratification by cytology and HPV genotype to inform the management of HPV-positive women in routine cervical screening. Br J Cancer 2020; 122 (11), 1715–23.

21. Zhao J, Guo Z, Wang Q, et al. Human papillomavirus genotypes associated with cervical precancerous lesions and cancer in the highest area of cervical cancer mortality, Longnan, China. Infect Agents Cancer. 2017; 12:8.

22. Namujju PB, Waterboer T, Banura C, et al. Risk of seropositivity to multiple oncogenic human papillomavirus types among human immunodeficiency virus-positive and -negative Ugandan women. J General Virol. 2011; 92 (12), 2776–83

23. Hanischa RA, Sowc PS, Tourec M, et al. Influence of HIV-1 and/or HIV-2 infection and CD4 count on cervical HPV DNA detection in women from Senegal, West Africa. J Clin Virol. 2013; 58 (4), 696–702.

24. Taku O, Businge CB, Mdaka ML, et al. Human papillomavirus prevalence and risk factors among HIV-negative and HIV-positive women residing in rural Eastern Cape, South Africa. Int. J Infect Dis. 2020; 95, 176–82

25. Huh WK, Joura EA, Giuliano AR et al. Final efficacy, immunogenicity, and safety analyses of a nine-valent human papillomavirus vaccine in women aged 16–26 years: a randomised, double-blind trial. The Lancet. 2017. Doi: 10.1016/S0140-6736(17)31821-4

26. Pirek D, Petignat P, Vassilakos P, et al. Human papillomavirus genotype distribution among Cameroonian women with invasive cervical cancer: a retrospective study. Sex Transm Infect. 2015; 91 (6), 440–44.

27. Njuguna DW, Mahrouseh N, Onisoyonivosekume D, Varga O. National Policies to Prevent and Manage Cervical Cancer in East African Countries: A Policy Mapping Analysis. Cancers 2020, 12 (6), 1520.

28. Obiri-Yeboah D, Akakpo PK, Mutocheluh M, et al. Epidemiology of cervical human papillomavirus (HPV) infection and squamous intraepithelial lesions (SIL) among a cohort of HIV-infected and uninfected Ghanaian women. BMC Cancer. 2017; 17 (1) 688

29. Yakub MM, Fowotade A, Anaedobe CG, et al. Human papillomavirus correlates of high grade cervical dysplasia among HIV-Infected women at a major treatment centre in Nigeria: a cross-sectional study. Pan Afr Med J. 2019; 33, 125.

30. Mutombo AB, Benoy I, Tozin R, Bogers J, Van geertruyden JP, Jacquemyn Y. Prevalence and Distribution of Human Papillomavirus Genotypes Among Women in Kinshasa, The Democratic REpublic of the Congo. J Global Oncol 2019; 1–9.

31. Ghedira R, Mahfoudh W, Hadhri S, et al. Human papillomavirus genotypes and HPV16 variants distribution among Tunisian women with normal cytology and squamous intraepithelial lesions. Infect Agents Cancer. 2016; 11 (1), 1–10.

32. Mchome BL, Kjaer SK, Manongi R, et al. HPV types, cervical high-grade lesions and risk factors for oncogenic human papillomavirus infection among 3416 Tanzanian women. Sex Transm Infect. 2020. doi:10.1136/sextrans-2019-054263

33. Vassilakos P, Catarino R, Bougel S, et al. Use of Swabs for dry collection of self-samples to detect human papillomavirus among Malagasy women. Infect Agents Cancer 2016; 11 (1), 13.

34. Marembo T, Mandishora RD, Borok M. Use of Multiplex Polymerase Chain Reaction for Detection of High-Risk Human Papillomavirus Genotypes in Women Attending Routine Cervical Cancer Screening in Harare. Intervirol 2019; 62 (2), 90–5. doi: 10.1159/000502206

35. Belglaiaa E, Elannaz H, Mouaouya B, et al. Human papillomavirus genotypes among women with or without HIV infection: an epidemiological study of Moroccan women from the Souss area. Infect Agents Cancer 2015; 10 (1), 1–10

36. Nyasenu YT, Gbeasor-Komlanvi FA, Issa SA, et al. Prevalence of HPV among HIV-negative women of child-bearing age in Lomé, Togo. Future Virol. 2019; 14 (12), 783–90

37. Menon SS, Rossi R, Harebottle R, Mabeya H, vanden Broeck D. Distribution of human papillomaviruses and bacterial vaginosis in HIV positive women with abnormal cytology in Mombasa, Kenya. Infect Agents Cancer. 2016; 11 (1), 17.

38. Krings A, Dunyo P, Pesic A, et al. Characterization of HumanPapillomavirus prevalence and risk factors to guide cervical cancer screening in the North Tongu District, Ghana. PLoS One. 2019; 14 (6): e0218762.

39. Ezechi OC, Ostergren PO, Nwaokorie FO, Ujah IA, Pettersson KO. The burden, distribution and risk factors for cervical oncogenic human papilloma virus infection in HIV positive Nigerian women. Virol J. 2014, 11 (1), 5

40. Youssef MA, Abdelsalam L, Harfoush RA, et al. Prevalence of human papilloma virus (HPV) and its genotypes in cervical specimens of Egyptian women by linear array HPV genotyping test. Infect Agents Cancer. 2016; 11 (1), 6

41. Awua AK, Severini A, Wiredu EK, Afari EA, Zubach VA, Adanu RM. Self-Collected Specimens Revealed a Higher Vaccine- and Non-Vaccine-Type Human Papillomavirus Prevalences in a Cross-Sectional Study in Akuse. Adv Prev Med. 2020. Doi: 10.1155/2020/8343169

42. Boumba LMA, Qmichou Z, Mouallif M, et al. Human Papillomavirus Genotypes Distribution by Cervical Cytologic Status among Women Attending the General Hospital of Loandjili, Pointe-Noire, Southwest Congo (Brazzaville). J Med Virol. 2015; 87:1769–76.

43. Akarolo-Anthony SN, Maryam Al-Mujtaba, Ayotunde O Famooto, Eileen O Dareng, Olayinka B Olaniyan, Richard Offiong, Cosette M Wheeler and Clement A Adebamowo. HIV associated high-risk HPV infection among Nigerian women. BMC Infect Dis. 2013; 13 (1), 1–6

44. Reddy D, Njala J, Stocker P, et al. High-risk human papillomavirus in HIV-infected women undergoing cervical cancer screening in Lilongwe, Malawi: a pilot study. International Journal of STD & AIDS 2015; 26 (6), 379–87

45. Sweet K, Bosire C, Sanusi B, et al. Prevalence, incidence, and distribution of human papillomavirus types in female sex workers in Kenya. Int J STD & AIDS 2020; 31(2) 109–118

46. Dols JA, Reid G, Brown JM, et al. HPV type distribution and cervical cytology among HIV-positive Tanzanian and South African women. ISRN Obstet Gynecol. 2012. doi:10.5402/2012/514146

47. Said HM, Ahmed K, Burnett R, Allan BR, Williamson AL, Hoosen AA. HPV genotypes in women with squamous intraepithelial lesions and normal cervixes participating in a community-based microbicide study in Pretoria, South Africa. J Clin Virol. 2009; 44 (4), 318–21

48. Wolday D, Derese M, Gebressellassie S, et al. HPV genotype distribution among women with normal and abnormal cervical cytology presenting in a tertiary gynecology referral Clinic in Ethiopia. Infect Agents Cancer. 2018; 13 (1), 28.

49. Maranga IO, Hampson L, Oliver AW, et al. HIV Infection Alters the Spectrum of HPV Subtypes Found in Cervical Smears and Carcinomas from Kenyan Women. The Open Virol J. 2013; 7, 19–27

50. Vidal AC, Murphy SK, Hernandez BY, et al. Distribution of HPV genotypes in cervical intraepithelial lesions and cervical cancer in Tanzanian women. Infect Agents Cancer. 2011, 6 (1), 20.

51. Guthrie BL, Rositch AF, Cooper JA, Farquhar C, Bosire R, Choi R, Kiarie J, Smith JS. Human papillomavirus and abnormal cervical lesions among HIV-infected women in HIV-discordant couples from Kenya. Sexually transmitted infections. 2020. Doi: 10.1136/sextrans-2019-054052

52. Okolo C, Franceschi S, Adewole I, et al. Human papillomavirus infection in women with and without cervical cancer in Ibadan, Nigeria. Infect Agents Cancer 2010, 5 (1), 24

53. Zhang C, Huang C, Zheng X, Pan D. Prevalence of human papillomavirus among Wenzhou women diagnosed with cervical intraepithelial neoplasia and cervical cancer. Infect Agents Cancer. 2018; 13 (2018), 37

54. Thapa N, Maharjan M, Shrestha G, et al. Prevalence and type-specific distribution of human papillomavirus infection among women in mid-western rural, Nepal-A population-based study. BMC Infect Dis. 2018; 18 (1), 338

55. Tan SC, Ismail MP, Duski DR, Othman NH, Ankathil R. Prevalence and type distribution of human papillomavirus (HPV) in Malaysian women with and without cervical cancer: an updated estimate. Biosci Rep. 2018; 38 (2), BSR20171268

56. Kesheh MM, Hossein Keyvani. The Prevalence of HPV Genotypes in Iranian Population: An Update. Iran J Pathol. 2019; 14 (3), 197–205

57. Van S, Khac MN, Dimberg J, Matussek A, Henningsson AJ. Prevalence of cervical infection and genotype distribution of human papilloma virus among females in Da Nang, Vietnam. Anticancer Res. 2017; 37 (3), 1243–7.

58. Li Z, Liu F, Cheng S, et al. Prevalence of HPV infection among 28,457 Chinese women in Yunnan Province, southwest China. Sci Rep. 2016; 6, 21039

59. Zhong TY, Zhou JC, Hu R, et al. Prevalence of human papillomavirus infection among 71,435 women in Jiangxi Province China. J Infect Public Health 2017; 10 (6), 783–88

60. Zhang J, Zhang D, Yang Z, Wang X, Wang D. The role of human papillomavirus genotyping for detecting high-grade intraepithelial neoplasia or cancer in HPVpositive women with normal cytology: a study from a hospital in northeastern China. BMC Cancer. 2020; 20, 443

61. Zeng Z, Yang H, Li Z, et al. Distribution of HPV Infection in China: Analysis of 51,345 HPV Genotyping Results from China’s Largest CAP Certified Laboratory. J Cancer. 2016; 7 (9), 1037

62. Bansal D, Elimi AA, Skariah S, et al. Molecular epidemiology and genotype distribution of Human papillomavirus (HPV) among Arab women in the state of Qatar. J Trans Med. 2014, 12 (1), 1–9

63. Wang R, Guo XL, Wisman GB, et al. Nationwide prevalence of human papillomavirus infection and viral genotype distribution in 37 cities in China. BMC Infect Dis. 2015; 15 (1), 257.

64. Kantathavorn N, Mahidol C, Sritana N. et al. Genotypic distribution human papillomavirus (HPV) and cervical cytology findings in 5906 Thai women undergoing cervical cancer screening programs. Infect Agents Cancer. 2015. Doi: 10.1186/s13027-015-0001-5

65. Park EK, Cho H, Lee SH, et al. Human Papillomavirus Prevalence and Genotype Distribution among HIV-Infected Women in Korea. J Korean Med Sci. 2014; 29 (1), 32–7

66. Miyashita M, Agdamag DM, Sasagawa T, et al. High-Risk HPV Types in Lesions of the Uterine Cervix of Female Commercial Sex Workers in the Philippines. J Med Virol. 2009; 81 (3), 545–51.

67. Vet JNI, de Boer MA, van den Akker BEWN, et al. Prevalence of human papillomavirus in Indonesia: a population-based study in three regions. Br J Cancer. 2008; 99 (1), 214–18

